# Engagement and factors associated with annual anal human papillomavirus screening among sexual and gender minority individuals

**DOI:** 10.1101/2024.04.22.24306185

**Authors:** Jenna Nitkowski, Timothy J. Ridolfi, Sarah J. Lundeen, Anna R. Giuliano, Elizabeth Chiao, Maria E. Fernandez, Vanessa Schick, Jennifer S. Smith, Paige Bruggink, Bridgett Brzezinski, Alan G. Nyitray

**Affiliations:** Center for AIDS Intervention Research, Medical College of Wisconsin, Milwaukee, Wisconsin, USA; Department of Surgery, Medical College of Wisconsin, Milwaukee, Wisconsin, USA; Center for Immunization and Infection Research in Cancer, Moffitt Cancer Center and Research Institute, Tampa, Florida, USA; MD Anderson Cancer Center, The University of Texas, Houston, Texas, USA; Department of Health Promotion and Behavioral Sciences, The University of Texas Health Science Center at Houston School of Public Health, Houston, Texas, USA; Department of Management, Policy and Community Health, The University of Texas Health Science Center at Houston School of Public Health, Houston, Texas, USA; Gillings School of Global Public Health, University of North Carolina at Chapel Hill, Chapel Hill, North Carolina, USA; Medical College of Wisconsin, Milwaukee, Wisconsin, USA; Clinical Cancer Center, Medical College of Wisconsin, Milwaukee, Wisconsin, USA

**Keywords:** anal cancer, sexual and gender minority individuals (SGM), human papillomavirus (HPV), self-sampling, screening

## Abstract

**Objectives:** Annual screening with a provider has been recommended for groups at highest risk for anal cancer. Anal self-sampling could help address screening barriers, yet no studies have examined annual engagement with this method.

**Methods:** The Prevent Anal Cancer Self-Swab Study recruited sexual and gender minority individuals 25 years and over who have sex with men in Milwaukee, Wisconsin to participate in an anal cancer screening study. Participants were randomized to a home or clinic arm. Home-based participants were mailed an anal human papillomavirus self-sampling kit at baseline and 12 months, while clinic-based participants were asked to schedule and attend one of five participating clinics at baseline and 12 months. Using Poisson regression, we conducted an intention-to-treat analysis of 240 randomized participants who were invited to screen at both timepoints.

**Results:** 58.8% of participants completed annual (median=370 days) anal screening. When stratified by HIV status, persons living with HIV had a higher proportion of home (71.1%) versus clinic (22.2%) annual screening (*p*<0.001). Non-Hispanic Black participants had a higher proportion of home-based annual anal screening engagement (73.1%) compared to annual clinic screening engagement (31.6%) (*p*=0.01). Overall, annual screening engagement was significantly higher among participants who had heard of anal cancer from an LGBTQ organization, reported “some” prior anal cancer knowledge, preferred an insertive anal sex position, and reported a prior cancer diagnosis. Annual screening engagement was significantly lower for participants reporting a medical condition.

**Conclusions:** Annual screening engagement among those at disproportionate anal cancer risk was higher in the home arm.

## Introduction

Anal cancer disproportionately affects men who have sex with men, particularly if they are living with HIV.(1) Men who have sex with men and trans women living with HIV age 45 and over have a 10-fold higher anal cancer incidence versus the general population.(1, 2) Recently, consensus guidelines for screening those at highest risk for anal cancer have been developed.(2) Although large variation in screening availability and resources inhibits a standardized screening strategy for all settings, initial screening typically consists of anal cytology and human papillomavirus (HPV) testing using clinician-collected anal swabs in addition to a digital anal rectal examination.(2) For negative test results, repeat screening at 12 months is advised.(2)

Home-based anal self-sampling is a potential method of anal swab collection to reach individuals for anal precancer screening. Research has shown high acceptability and willingness to use an anal self-sampling test among sexual minority men. (3–5) In the Prevent Anal Cancer Self-Swab Study, a randomized controlled trial which compared home versus clinic anal swabbing,(6) participants were more likely to return a mailed home-based swab than attend a clinic for swabbing by a clinician.(7) Home-based anal screening engagement was especially higher among persons living with HIV and Black individuals, two groups disproportionately affected by anal cancer.(7)

To our knowledge, no studies have examined engagement with annual anal HPV DNA self-sampling. The International Anal Neoplasia Society (IANS) anal cancer screening consensus guidelines pertain to clinic-based screening, so home collection testing and intervals have not yet been established. But given that repeat testing is advised for one year later for negative results, it is likely that screening either at home or in a clinic would be annually. In this research, we present results of engagement with mailed annual home-based anal canal HPV DNA self-sampling and annual clinic-based anal HPV DNA swabbing. We also assess factors associated with annual swabbing.

## Methods

### Study design and recruitment

The Prevent Anal Cancer Self-Swab Study is a randomized controlled trial which examined anal cancer screening among sexual and gender minority individuals in the Milwaukee, Wisconsin area between 2020 and 2023. The protocol has previously been published.(6) Participants were recruited via social media ads, flyers distributed in targeted businesses and non-profit organizations, community events, and a voluntary referral program. Eligibility criteria consisted of the following: 25 years of age or over, assigned male sex at birth or transgender gender identity, acknowledgement of sex with men in the last five years or identification as gay or bisexual, and willingness to provide informed consent and comply with the study protocol. Individuals who planned to move within 12 months, reported unwillingness to attend one of the designated study clinics, had a prior diagnosis of anal cancer, or reported use of anticoagulants other than aspirin or non-steroidal anti-inflammatory drugs were excluded. The study activities were approved by the Medical College of Wisconsin Human Protections Committee.

After providing informed consent, participants were asked to complete a baseline computer-assisted self-interview (CASI) survey. The baseline survey contained questions about attitudes, behaviors, and prior healthcare procedures. Participants were then randomized in a one-to-one allocation to either a home-based or clinic-based arm. The overall objective of the study was to evaluate engagement with annual home versus clinic anal HPV DNA screening among sexual and gender minority individuals. Home-based participants were sent an anal self-sampling kit in the mail at baseline and one year later to complete and return via mail. Clinic-based participants were asked to attend a clinic appointment at baseline and at 12 months where a clinician collected an anal swab. During the consenting session, participants were told that they will be asked to repeat the procedures one year later so no recommendation about annual screening was given by study staff or providers. All participants were asked to attend an initial clinic appointment for a digital anal rectal examination followed by a subsequent appointment at 12 months for a high-resolution anoscopy (HRA).

#### Home arm

Participants randomized to the home-based arm were mailed an anal HPV DNA self-sampling kit. The kit consisted of a flocked swab (COPAN Italia S.p.A., Brescia, Italy), a vial of standard transport medium (QIAGEN, Germantown, MD, USA), a pair of gloves, illustrated instructions written at a sixth-grade reading level in English or Spanish modeled after published instructions,(8) and instructions and packaging for returning the swab. An identical kit was mailed 12 months later. Regardless of whether a participant returned the baseline kit, all participants in the home-based arm were mailed a 12-month kit. Study staff contacted participants who did not return the kit via their preferred contact method up to three times.

#### Clinic arm

Participants in the clinic arm were asked to attend an appointment at their choice of five area clinics. To mimic real-world conditions, participants were asked to call and schedule their clinic appointment. At the clinic appointment, biometric measurements including height, weight, and waist circumference were taken. A highly experienced clinician then collected an anal swab from the participant and performed a digital anal rectal examination. The swabs, the standard transport medium, and the swabbing technique instructions were identical in both the home and clinic. Participants were contacted prior to their 12-month anniversary date to schedule the follow-up appointment regardless of attendance at the baseline appointment. Because the 12-month clinic appointment and HRA were designed to occur one year after the baseline clinic appointment, some clinic participants opted to combine them. Out of the 71 clinic participants who completed both a 12-month clinic appointment and HRA, 71.8% (n=51) combined these appointments.

Between January 2020 and August 2022, a total of 240 participants were randomized to the study (home=120, clinic=120). Study activities were paused between March 2020 and November 2020 due to the COVID-19 pandemic.

### Measures

#### Outcome

Annual anal screening engagement was the outcome. This was coded as a dichotomous variable (1=yes, 0=no) representing whether a participant provided an anal swab at both baseline and 12 months. For home participants, this meant returning both a baseline and 12-month kit. For clinic participants, this meant attending a baseline clinic appointment and 12-month clinic appointment/HRA.

#### Exposures

The primary exposure of interest was study arm (home-based or clinic-based anal screening). Participant characteristics, behaviors, attitudes, and prior health care procedures were all examined as possible exposures. These variables come from the eligibility and baseline participant surveys and consist of age, race/ethnicity, education, marital status, HIV status, preferred anal sex position, sources of anal cancer awareness, amount of prior anal cancer knowledge, fear of screening, history of a cancer diagnosis, history of doing a self-examination, history of anal cancer screening procedures such as anal cytology, smoking status, and ever being diagnosed with certain medical conditions. We also calculated the 2021 area deprivation index (ADI) score for each participant’s address using the Neighborhood Atlas mapping function. (9)

For sources of anal cancer awareness, participants were asked “*Where have you heard about anal cancer before today? (check all that apply)*”. Response options included: doctor’s office, community health worker, public health department, school or university, hospital or research institution, friend or acquaintance, LGBTQ community organization, online, etc. For the medical condition question, participants were asked “*Here is a list of medical conditions that may make it harder to use the swab. Has a doctor ever said that you have any of the following? (check all that apply)*.” Response options consisted of the following: arthritis (n=26), carpal tunnel syndrome (n=7), cerebral palsy (n=1), deafness (n=4), diabetes (n=16), fibromyalgia (n=1), chronic lower back pain (n=19), motor neuron diseases (n=1), movement disorders (n=4), multiple sclerosis (n=1), obesity (n=34), spina bifida (n=1), spinal cord injury (n=4), stroke (n=2), visual impairment (n=10), other medical condition (n=3), or none of the above (n=151). Responses were combined to create a dichotomous variable representing whether a participant reported any medical condition (1=yes, 0=no).

### Statistical analysis

We conducted an intention-to-treat analysis to evaluate engagement and factors associated with annual home and clinic anal HPV DNA screening (n=240). Univariate and multivariable Poisson regression analyses were conducted to evaluate factors associated with annual anal screening engagement. Factors with a p-value of less than 0.25 in bivariate analyses were included in a multivariable regression model. Manual backward elimination was used to remove factors with a p-value greater than 0.05 until the remaining factors had a p-value less than or equal to 0.05. Potential confounders were included in the final model. Missing data were handled by pairwise deletion. Response option categories were combined in instances of small cell sizes. Relative risk was calculated using Poisson regression with robust standard errors and the log-link function in Stata. We report unadjusted and adjusted relative risk ratios with 95% confidence intervals. All analyses were conducted in SPSS 28 (Armonk, NY, USA) and Stata BE 18 (College Station, TX, USA).

## Results

Overall, 58.8% (n=141) of 240 randomized participants engaged in annual anal HPV screening with a median of 370 days between screenings. In the home arm, 65.0% of home-based participants engaged in annual screening compared to 52.5% of clinic-based participants (*p*=0.049) (Figure 1).

**Figure 1.**
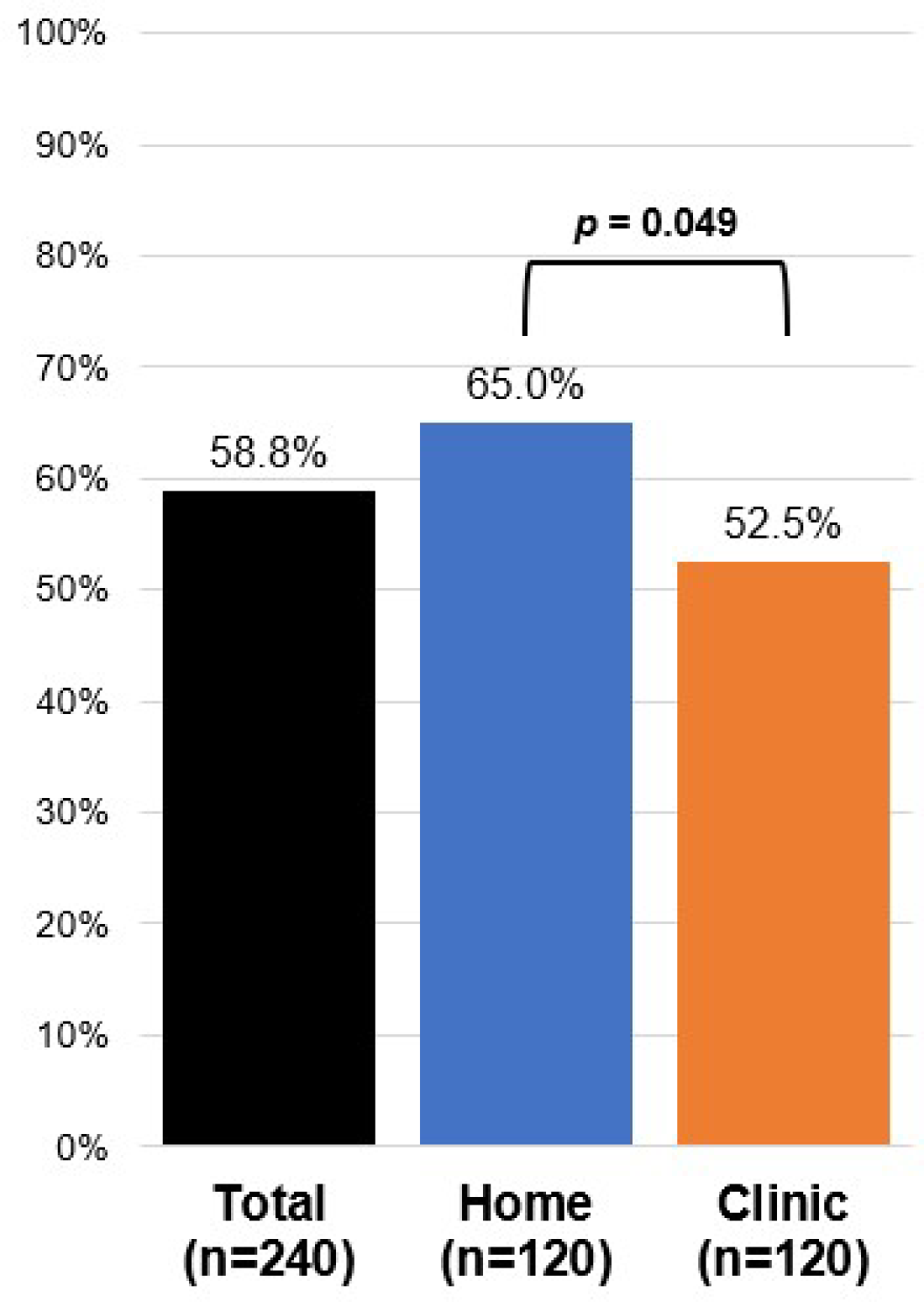
Proportion of participants who engaged in annual anal HPV sampling in the Prevent Anal Cancer Self-Swab Study, Milwaukee, Wisconsin, 2020-2023.

Table 1 presents the characteristics of randomized participants stratified by annual screening engagement. The average age was 46 years and ranged from 25 to 78 years. Most participants (94.6%) identified as a man, 2.9% as transgender, and 2.5% as non-binary or other. Nearly two-thirds of participants (66.1%) identified as non-Hispanic White, 18.8% as non-Hispanic Black, 13.0% as Hispanic or Latino/x, and 2.1% as non-Hispanic Other. More than one in four participants (27.1%) were living with HIV, and most reported never or not knowing if they had ever had an anal cytology test (75.8%) and little or no prior anal cancer knowledge (78.8%).

**Table 1.**
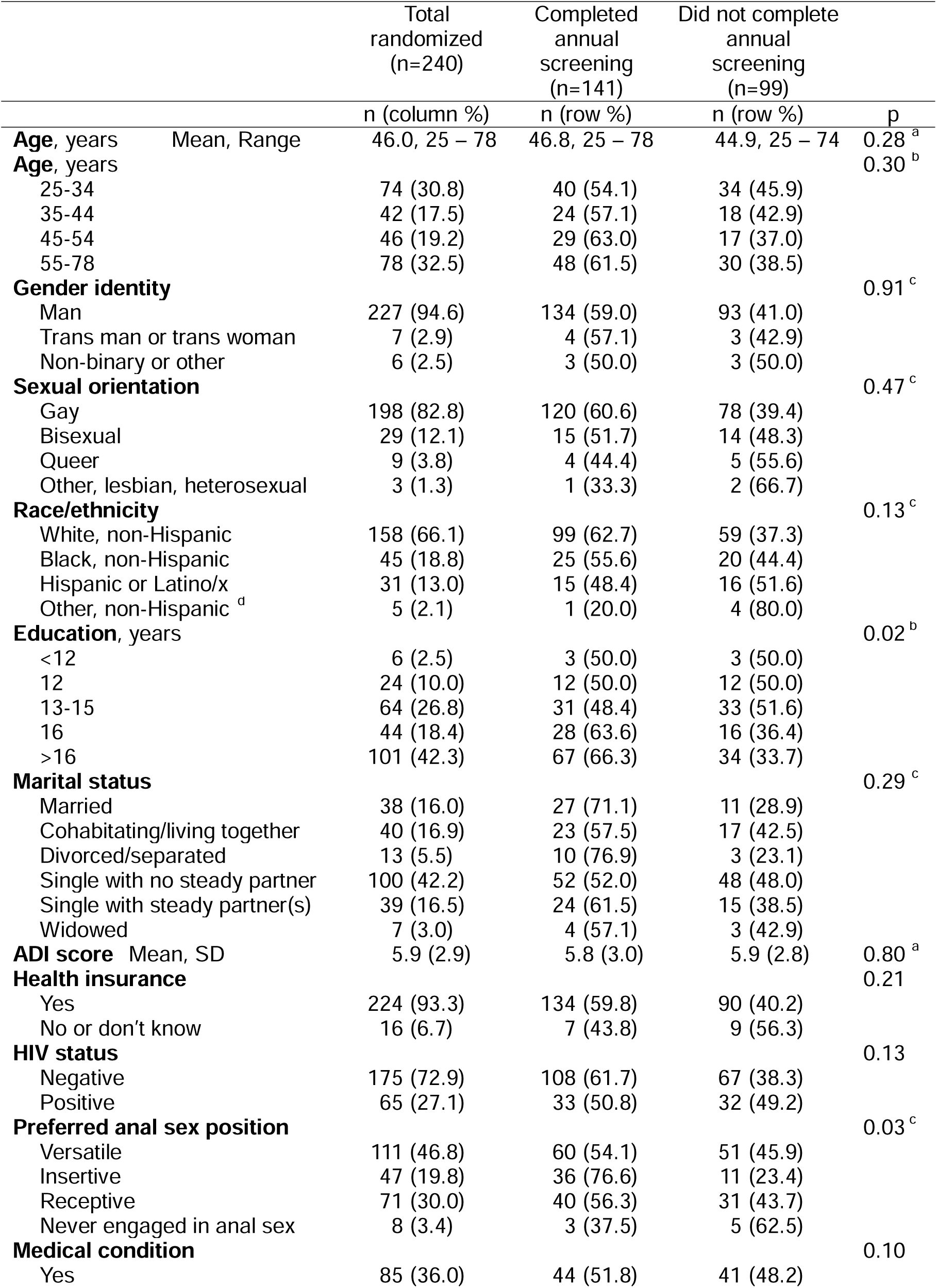
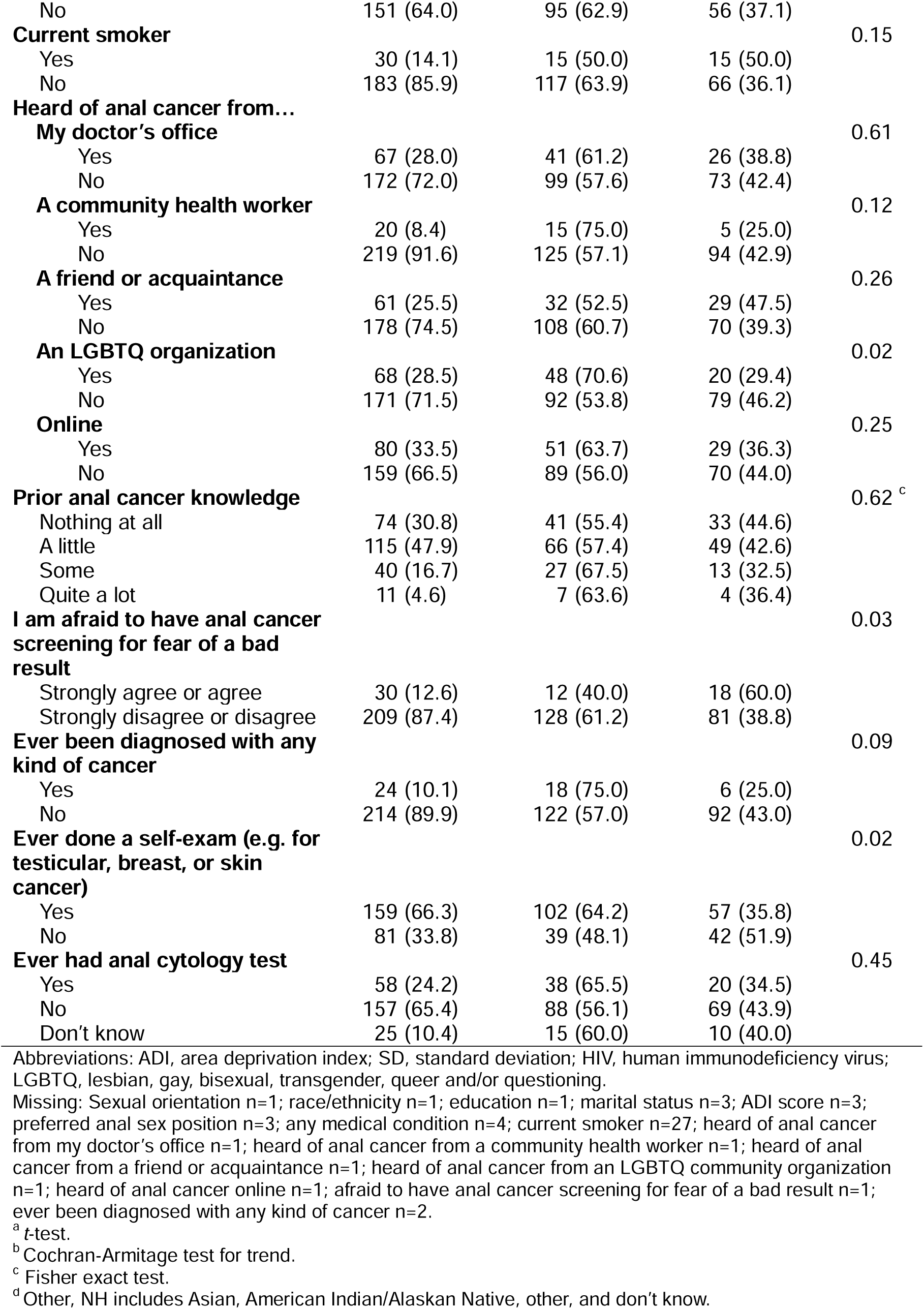
Intention-to-treat analysis of randomized participants who completed annual anal human papillomavirus swabbing in the Prevent Anal Cancer Self-Swab Study, Milwaukee, Wisconsin, USA, 2020-2023.

When stratifying by race/ethnicity (Figure 2), non-Hispanic Black participants had a higher proportion of home-based annual anal screening engagement (73.1%) compared to annual clinic screening engagement (31.6%) (*p*=0.01). When stratifying by HIV status (Figure 2), persons living with HIV had a higher proportion of home-based annual anal screening engagement (71.1%) compared to clinic screening engagement (22.2%) (*p*<0.001). In contrast, HIV-negative participants had nearly equal proportions of engaging in home (62.2%) versus clinic (61.3%) annual screening (*p*=0.90).

**Figure 2.**
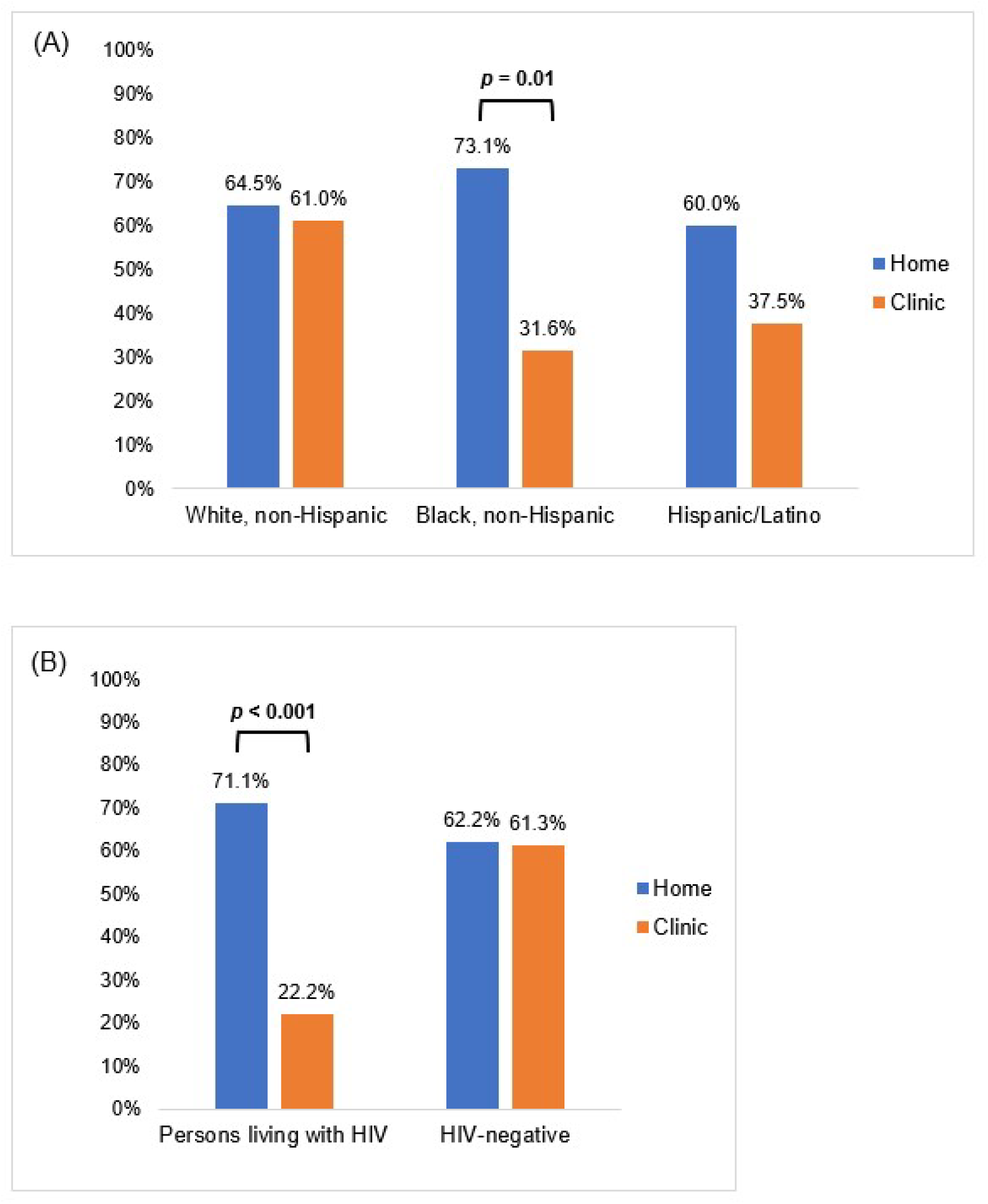
Proportion of participants who engaged in annual anal HPV sampling by study arm stratified by (A) race/ethnicity and (B) HIV status in the Prevent Anal Cancer Self-Swab Study, Milwaukee, Wisconsin, 2020-2023.

Results of univariate and multivariable Poisson regression analyses are presented in Table 2. Although not significant, participants in the home-based arm were more likely to engage in annual anal HPV screening (aRR 1.20, 95% CI 0.97 – 1.48) than the clinic arm while controlling for participant characteristics. Participants who reported ever being diagnosed with a medical condition were significantly less likely to engage in annual screening (aRR 0.78, 95% CI 0.61 – 0.998) compared to those who did not report a medical condition.

**Table 2.**
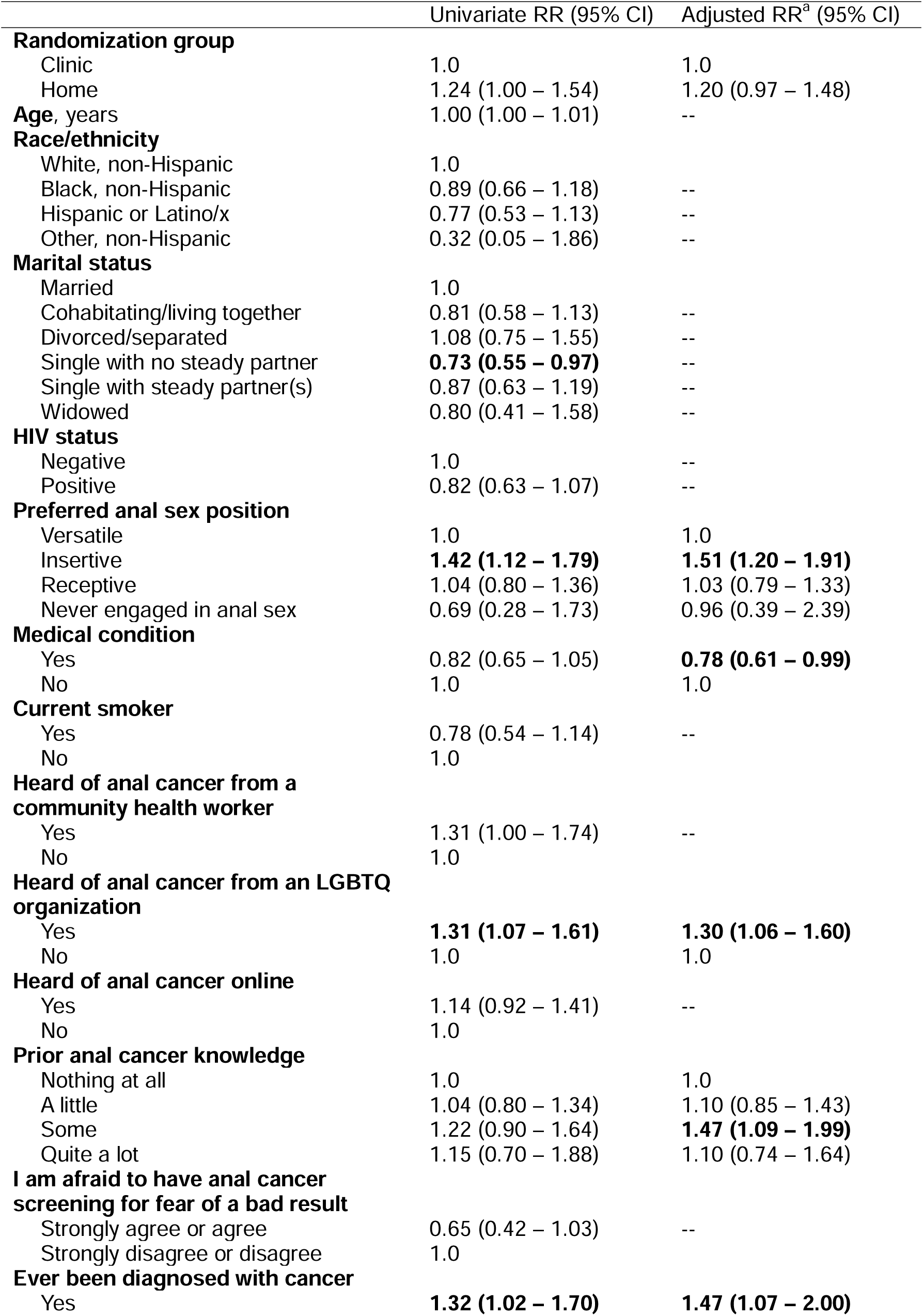
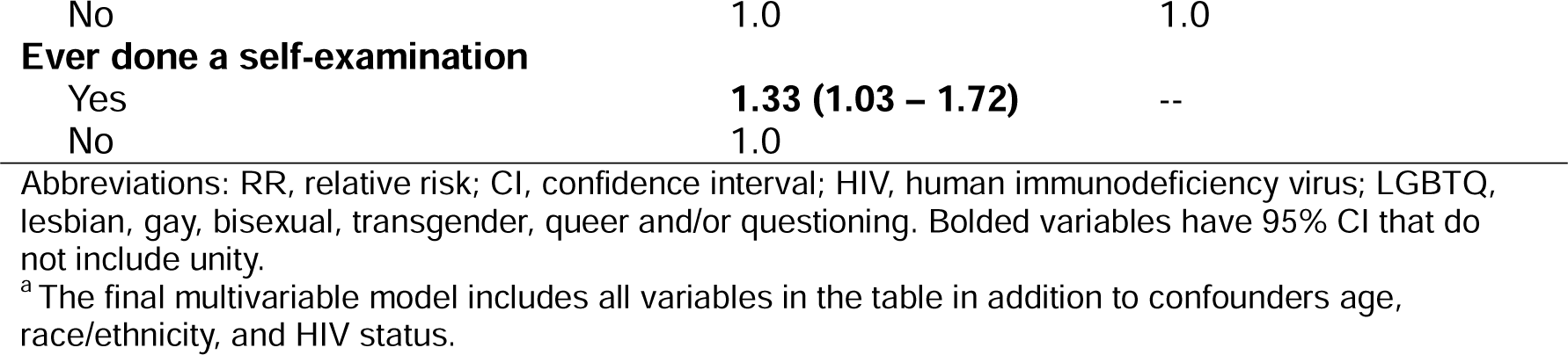
Factors associated with annual anal HPV screening engagement in the Prevent Anal Cancer Self-Swab Study, Milwaukee, Wisconsin, 2020-2023 (n=240).

|  | Univariate RR (95% CI) | Adjusted RR<sup>a</sup> (95% CI) |
| --- | --- | --- |
| **Randomization group** |  |  |
| Clinic | 1.0 | 1.0 |
| Home | 1.24 (1.00 – 1.54) | 1.20 (0.97 – 1.48) |
| **Age, years** | 1.00 (1.00 – 1.01) | -- |
| **Race/ethnicity** |  |  |
| White, non-Hispanic | 1.0 |  |
| Black, non-Hispanic | 0.89 (0.66 – 1.18) | -- |
| Hispanic or Latino/x | 0.77 (0.53 – 1.13) | -- |
| Other, non-Hispanic | 0.32 (0.05 – 1.86) | -- |
| **Marital status** |  |  |
| Married | 1.0 |  |
| Cohabiting/living together | 0.81 (0.58 – 1.13) | -- |
| Divorced/separated | 1.08 (0.75 – 1.55) | -- |
| Single with no steady partner | **0.73 (0.55 – 0.97)** | -- |
| Single with steady partner(s) | 0.87 (0.63 – 1.19) | -- |
| Widowed | 0.80 (0.41 – 1.58) | -- |
| **HIV status** |  |  |
| Negative | 1.0 | -- |
| Positive | 0.82 (0.63 – 1.07) | -- |
| **Preferred anal sex position** |  |  |
| Versatile | 1.0 | 1.0 |
| Insertive | **1.42 (1.12 – 1.79)** | **1.51 (1.20 – 1.91)** |
| Receptive | 1.04 (0.80 – 1.36) | 1.03 (0.79 – 1.33) |
| Never engaged in anal sex | 0.69 (0.28 – 1.73) | 0.96 (0.39 – 2.39) |
| **Medical condition** |  |  |
| Yes | 0.82 (0.65 – 1.05) | **0.78 (0.61 – 0.99)** |
| No | 1.0 | 1.0 |
| **Current smoker** |  |  |
| Yes | 0.78 (0.54 – 1.14) | -- |
| No | 1.0 |  |
| **Heard of anal cancer from a community health worker** |  |  |
| Yes | 1.31 (1.00 – 1.74) | -- |
| No | 1.0 |  |
| **Heard of anal cancer from an LGBTQ organization** |  |  |
| Yes | **1.31 (1.07 – 1.61)** | **1.30 (1.06 – 1.60)** |
| No | 1.0 | 1.0 |
| **Heard of anal cancer online** |  |  |
| Yes | 1.14 (0.92 – 1.41) | -- |
| No | 1.0 |  |
| **Prior anal cancer knowledge** |  |  |
| Nothing at all | 1.0 | 1.0 |
| A little | 1.04 (0.80 – 1.34) | 1.10 (0.85 – 1.43) |
| Some | 1.22 (0.90 – 1.64) | **1.47 (1.09 – 1.99)** |
| Quite a lot | 1.15 (0.70 – 1.88) | 1.10 (0.74 – 1.64) |
| **I am afraid to have anal cancer screening for fear of a bad result** |  |  |
| Strongly agree or agree | 0.65 (0.42 – 1.03) | -- |
| Strongly disagree or disagree | 1.0 |  |
| **Ever been diagnosed with cancer** |  |  |
| Yes | **1.32 (1.02 – 1.70)** | **1.47 (1.07 – 2.00)** |
| No | 1.0 | 1.0 |
| **Ever done a self-examination** |  |  |
| Yes | **1.33 (1.03 – 1.72)** | -- |
| No | 1.0 |  |

Abbreviations: RR, relative risk; CI, confidence interval; HIV, human immunodeficiency virus; LGBTQ, lesbian, gay, bisexual, transgender, queer and/or questioning. Bolded variables have 95% CI that do not include unity.
<sup>a</sup>The final multivariable model includes all variables in the table in addition to confounders age, race/ethnicity, and HIV status.

Participants who reported a preferred insertive anal sex position (aRR 1.51, 95% CI 1.20 – 1.91) were significantly more likely to engage in annual screening compared to versatile participants. Those who reported hearing of anal cancer from an LGBTQ organization, reported some prior anal cancer knowledge, and reported a prior cancer diagnosis were significantly more likely to engage in annual screening. Participants who reported previously hearing about anal cancer from an LGBTQ organization were more likely to engage in annual screening (aRR 1.30, 95% CI 1.06 – 1.60) compared to those who had not. Those who reported some prior anal cancer knowledge were more likely to screen annually compared to those who had no prior anal cancer knowledge (aRR 1.47, 95% CI 1.09 – 1.99). Participants who had any previous cancer diagnosis were more likely to engage in annual screening (aRR 1.47 95% CI 1.07 – 2.00) than those who never had a cancer diagnosis.

## Discussion

Overall, more than half of randomized participants engaged in annual anal HPV screening. Building upon our previous findings indicating higher engagement in home-based anal screening among non-Hispanic Black individuals and those living with HIV at baseline,(7) our current data demonstrate similar findings for annual screening. When stratifying by race/ethnicity, non-Hispanic Black participants had over twice the proportion of engaging in home-based annual screening (73.1%) versus clinic-based annual screening (31.6%) (*p=*0.01). Since Black sexual minority men are less likely to report clinic-based anal cancer screening,(10) home sampling may be a way to increase screening engagement. When stratified by HIV status, 71.1% of persons living with HIV completed annual home-based anal screening compared to just 22.2% who engaged in annual clinic screening (*p*<0.001). One possible explanation for this finding is appointment fatigue, which individuals living with HIV may experience due to repeated tests and procedures.(11)

We also found that those who had ever been diagnosed with a medical condition were less likely to engage in annual screening, either in the home or clinic. This suggests that managing a chronic condition may be a barrier to screening engagement. Thus, mailed home-based anal self-sampling may be a potential way to ease the burden of managing a chronic condition and increase screening engagement.

Participants who had ever done a self-exam (such as for testicular, breast, or skin cancer) were significantly more likely to engage in annual screening in univariate but not multivariable analysis, due to adjusting for factors like prior cancer diagnosis as well as prior anal cancer knowledge and sources.

Anal cancer knowledge and information sources were significantly associated with higher annual screening engagement. Participants who reported some amount of prior anal cancer knowledge and those who reported hearing about anal cancer from an LGBTQ organization were more likely to engage in annual screening. This suggests that LGBTQ organizations may be important channels to increase knowledge and awareness of anal cancer among sexual and gender minority individuals. Notably, knowledge among gay and bisexual men that HPV causes anal cancer is low (12).

Campaigns to increase anal cancer knowledge and annual screening engagement may partner with LGBTQ organizations as an important source of information. Although provider recommendation is strongly associated with increased cancer screening uptake (13, 14), including anal cancer (15), we found no association between annual screening and hearing about anal cancer from a doctor’s office. However, hearing about anal cancer in a doctor’s office is different than receiving a doctor’s recommendation for screening.

Participants who reported a preference for a primarily insertive anal sex position were less likely to engage in annual anal screening compared to participants who reported a preference for versatile anal sex. While the reasons for this finding are unclear and may warrant future research, it may be due to stigma associated with being a receptive anal sex partner or “bottom” (16, 17). Gay and bisexual men who report less anal sex stigma have greater comfort with discussing anal sex with a health care provider and are more likely to engage in anal screening.(18) No significant differences in annual screening engagement were observed between cisgender men and non-cis gender persons, although only 13 non-cisgender participants were randomized to the study.

### Limitations

This research consisted of participants who chose to enroll in a randomized clinical trial about anal cancer screening. These individuals may differ from those who did not or would not choose to participate in this type of study. Thus, the findings from this research may not be representative of all sexual minority men or trans persons. It is also important to note that participants were not *encouraged* by study staff or study health care providers to attend annual screening. Participants were simply told by study staff during the consenting session that they will be asked to repeat the procedures one year later. Consensus screening guidelines were also released after the study ended.

While we asked participants whether they had received anal cancer screening procedures such as anal cytology and high-resolution anoscopy, we were not able to assess the quality of those procedures. Previous research has shown that having a negative healthcare experience can affect future healthcare engagement.(19) Finally, the medical condition question asked whether participants had *ever* been diagnosed so it does not capture whether they currently still have that condition. Future research with larger sample sizes of those with medical conditions can hopefully refine this finding.

## Conclusion

This study evaluated annual anal HPV screening engagement in the home and clinic. We found higher annual engagement with mailed home-based self-sampling compared to clinician swabbing, particularly among those living with HIV and non-Hispanic Black participants. Anal cancer education and its sources, such as LGBTQ organizations, may also play a role in increasing annual screening engagement. These findings underscore the importance of targeted educational efforts and provision of accessible screening methods in increasing anal cancer screening engagement, particularly among groups disproportionately affected by anal cancer.

## Acknowledgements

Thank you to the study participants, the Prevent Anal Cancer community advisory board, and the PAC Study Team (Cameron Liebert, Christopher Ajala, Madison Humphry, Esmeralda Lezama-Ruiz, Maritza Pallo, and Lisa Rein). Thanks to the clinicians who participated in this study: Mary Kay Schuknecht, Kathryn Kerhin, Nickie Gerboth, Dave Wenten, Annie Lakatos, Andrew Petroll, Brian Hilgeman, Leslie Cockerham, Sol del Mar Aldrete Audiffred, Winsome Panton, Christine Hogan, and Janaki Shah. We are grateful to the University of Chicago for help with the anal self-sampling illustrated instructions and to COPAN Italia S.p.A. for donation of some of the swabs used in this study.

## Data Availability Statement

Fully de-identified datasets and data dictionary will be shared with properly trained investigators on the study website within 1 year of study completion after assessment of institutional policies, Medical College of Wisconsin Human Research Protections Program rules, and local, state, and federal laws and regulations.

## Ethics Approval and Patient Consent

Informed consent was obtained from all study participants and study activities were approved by the Medical College of Wisconsin Human Research Protections Committee (protocol #PRO00032999).

## Declaration of generative AI and AI-assisted technologies in the writing process

The authors declare no generative AI or AI-assisted technologies in the writing process.

